# Assessing COVID-19 Pandemic Risk Perception and Response Preparedness in Veterinary and Animal Care Workers

**DOI:** 10.1101/2021.05.04.21256626

**Authors:** Kathryn R. Dalton, Kimberly M. Guyer, Francesca Schiaffino, Cusi Ferradas, Jacqueline R. Falke, Erin A. Beasley, Kayla Meza, Paige Laughlin, Jacqueline Agnew, Daniel J. Barnett, Jennifer B. Nuzzo, Meghan F. Davis

## Abstract

Veterinary and animal care workers (VACW) perform critical functions in biosecurity and public health, yet little has been done to understand the unique needs and barriers these workers face when responding during a pandemic crisis. This study evaluated VACWs’ perceived risks and roles during COVID-19, and explored barriers and facilitators in their readiness, ability, and willingness to respond during a pandemic. We deployed a survey targeting U.S. veterinary medical personnel, animal shelter and control workers, zoo and wildlife workers, plus other animal care workers. Data were collected on participants’ self-reported job and demographic factors, perceptions of risk and job efficacy, and readiness, ability, and willingness to respond during the pandemic. We found that leadership roles and older age had the strongest association with decreased perceived risk and improved job efficacy and confidence, and that increased reported contact level with others (both co-workers and the public) was associated with increased perceived risk. We determined that older age and serving in leadership positions were associated with improved readiness, willingness, and ability to respond. VACWs’ dedication to public health response, reflected in our findings, will be imperative if more zoonotic vectors of SARS-CoV-2 arise. Response preparedness in VACW can be improved by targeting younger workers not in leadership roles in support programs that center on improving job efficacy and confidence in safety protocols. These findings can be used to target intervention and training efforts to support the most vulnerable within this critical, yet often overlooked, workforce.

## Introduction

The 2019 coronavirus (SARS-CoV-2) pandemic, termed COVID-19, has caused extensive detrimental worldwide impacts. In the United States, there have been over 500,000 fatalities as of March 2021, one year since it was declared a national emergency.^1^ In times of crisis, the veterinary and animal care workforce is a source of unique knowledge and skills essential to national biosecurity and biological risk assessment and response.^2^ One of the functions of veterinary medicine and animal care workers (VACW) is to control infections within animal populations, including those that can transmit to people, therefore serving as a first line of defense against zoonotic diseases.^3^ In addition, they provide expert guidance on public health issues for their clients, visitors, and the public at large, such as communicating risks from exposure to pets.^3^ In addition to this distinctive role, VACW are a part of human response programs due to their knowledge of comparative medicine and public health (which is part of the veterinary oath and training); they have been employed in COVID-19 response efforts from human vaccination clinics to donation of medical supplies.^4,5^ Yet, VACW themselves are at risk for exposure to diseases from human-to-human transmission pathways that exist during normal business operations, and may also be exposured to zoonotic agents from animal patients. VACW may be reluctant to respond to work during a pandemic if they perceive it could put themselves and their families at risk.

To identify underlying causes of potential reluctance and the likelihood of response to work among VACW during a pandemic, we utilized the “Ready, Willing, and Able” model to characterize response preparedness.^6^ These components are differentiated by *Readiness* relating to the external infrastructure of personnel and material resources necessary to perform a task, *Willingness* as the predilection and desire to perform a task, and *Ability* referring to the skills and knowledge needed to actually perform the task. Research using the preparedness Ready, Willing, and Able model has been conducted on public health workers, healthcare employees, first responders, and other vital occupations for natural and biological disasters to capture factors that influence response preparedness.^7–14^ However, there are no data on risks and needs within the veterinary and animal care professions. No systematic research has been done to understand individual perceptions of needs, barriers, and facilitators related to working during a pandemic crisis in VACW, despite their importance to disaster preparedness and response. The current COVID-19 pandemic, and its profound ramifications for public health, provides a rationale for the importance of disaster preparedness and a resilient workforce. It can also serve as a natural experiment to address preparedness and response for this pandemic, as well as future biological and natural crises.

As such, the objectives of this research were to assess VACWs’ perceived risk of COVID-19 and their perception of their roles during this crisis. We explored barriers and facilitators to their readiness, willingness, and ability to respond during a crisis and determined factors that affected these outcomes. The findings from this research can be used to address risk communication needs, and to design and implement response interventions, including preparedness training and support systems for this vulnerable yet critical worker population. The ultimate goal of this research is to build an animal care workforce that is not only capable, but willing to respond during this and future crises.

## Methods

Approval for this study was received from The Johns Hopkins Bloomberg School of Public Heath Institutional Review Board (JHSPH IRB). As primary data collection for the survey instrument was anonymous, written consent was not required by JHSPH IRB. Nonetheless, all participants were provided an electronic disclosure statement describing the study and emphasizing voluntary participation and agreed to participate prior to beginning the survey.

### Data Collection

We targeted adult (>18 years old) animal care worker populations in the United States. This encompassed veterinary medical personnel – comprising veterinarians, veterinary technicians, veterinary assistants, hospital managers and other animal hospital support staff – who serve any patient type, including companion (e.g., dogs and cats), equine, laboratory, exotic, and food animals. Other target populations were animal shelter and animal control employees, laboratory animal personnel, zoological and wildlife facility workers, and those who self-identified in animal-related workforces, such as industry, government/advocacy, and academic research. We recruited study participants via email or phone through state licensing agencies, professional organizations, professional conference attendees, professional group listservs, social media, and existing contacts.

Survey data were collected and managed using REDCap electronic data capture tools hosted at Johns Hopkins Bloomberg School of Public Health.^15^ Survey data were collected from July 6, 2020 until Oct 25, 2020. The survey was an anonymous online questionnaire that consisted of two main sections: a demographic section and an attitude/belief section that focused on workers’ perceptions on their risk of exposure to COVID-19, the role they play in response efforts, and their response preparedness. Questions were developed based on feedback from experts and leaders within our target populations. Demographic and professional information included job title and role, years of employment, contact level with co-workers and the public (clients and visitors), geographic region, gender, age, race, marital status, household dependents, and annual household income. Key job and demographic questions can be found in ***Table 1, Table 2***, and ***Supplemental Table 1***. Because the survey was anonymous and participants were able to access it multiple times, we included data only from participants who completed all sections of the survey to minimize duplicate entries, although participants were allowed to skip questions within each section.

**Table 1:**
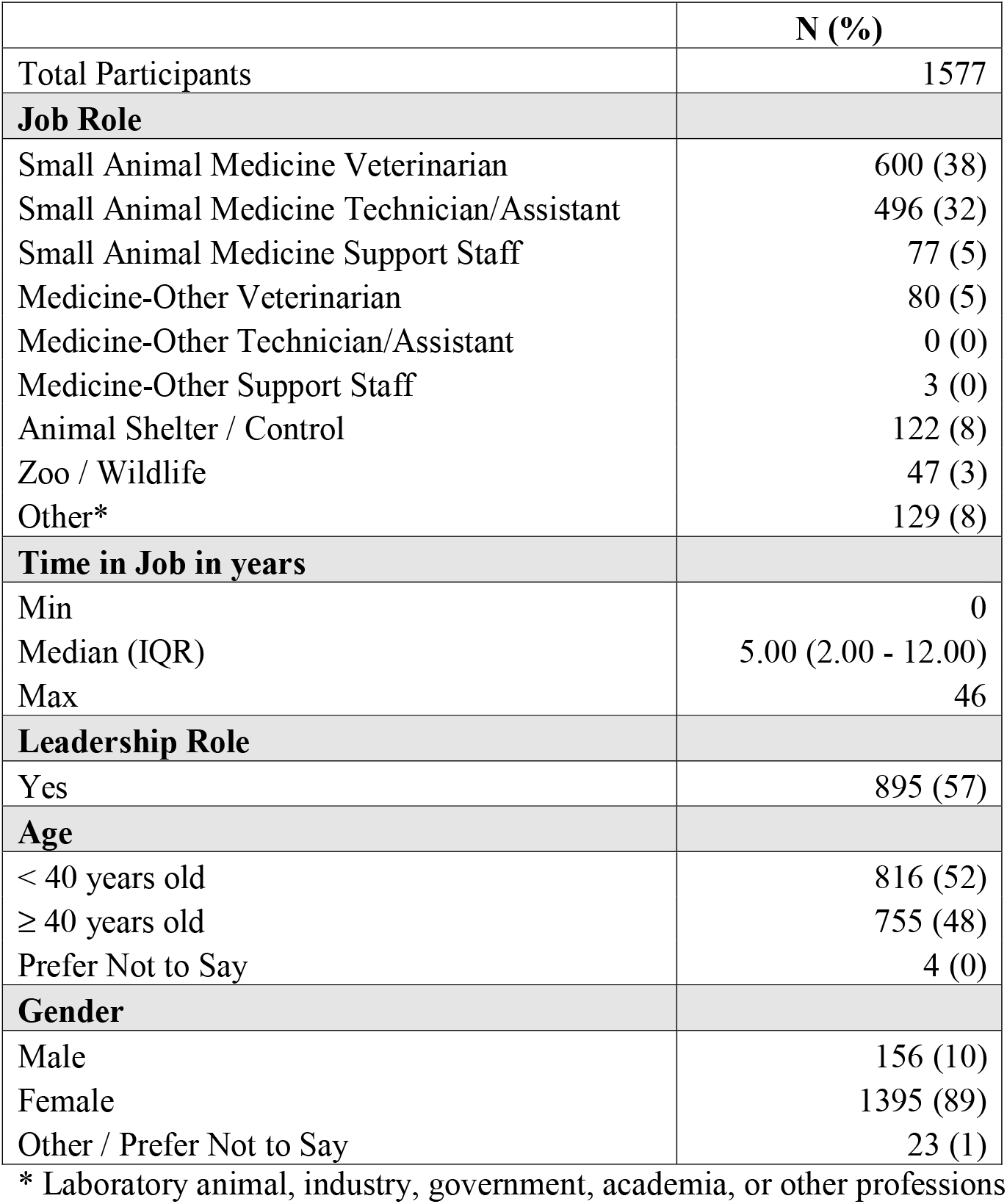
Job & Demographic Characteristics

**Table 2:** Contact Level Distribution

|  | **Co-Workers, N (%)** | **Public, N (%)** |
| --- | --- | --- |
| **Average Daily Contact** |  |  |
| No contact | 53 (3) | 295 (19) |
| Rarely (1-15%) | 89 (6) | 669 (43) |
| Intermittent (16-50%) | 181 (12) | 355 (23) |
| Most of the workday (50-85%) | 267 (17) | 174 (11) |
| Almost the entire workday (85-100%) | 983 (62) | 81 (5) |
| **Total People Contacted Daily** |  |  |
| 1-2 people | 99 (7) | 334 (26) |
| 3-5 people | 362 (24) | 233 (18) |
| 6-10 people | 594 (39) | 200 (16) |
| 11-24 people | 319 (21) | 267 (21) |
| 25+ people | 132 (9) | 227 (18) |
| **Contact Aggregated Score Quantile\*** |  |  |
| Minimal Contact (Q1) | 454 (31) | 374 (30) |
| Low Contact (Q2) | 362 (24) | 343 (27) |
| Moderate Contact (Q3) | 424 (29) | 235 (19) |
| High Contact (Q4) | 247 (17) | 296 (24) |

\* Sum of average daily contact and total people contacted daily, as ranked categorical variables (score 0-5), then split at 25%, 50%, and 75%.

Participants then answered questions regarding their attitudes and beliefs on their knowledge of the pandemic, confidence regarding safety protocols, perceived threat, job efficacy, likelihood of response barriers, and readiness, willingness, and ability to respond to the COVID-19 pandemic. We presented questions on a 5-point Likert scale, with a response of ‘5’ indicating strong agreement with the statement and a response of ‘1’ indicating strong disagreement. The distribution of responses is provided in ***Supplemental Table 2***. Additional questions included perceived primary sources of SARS-CoV-2 exposure, perceived consequences from the COVID-19 pandemic, and reported personal and professional barriers to working during the pandemic.

### Data Analysis

To assess potential risk to workers from contact with co-workers, clients, and the public, an aggregated contact level score was created by summing responses to the questions related to average daily contact and total people contacted and dividing the results into quartiles (minimal, low, moderate, and high contact).

We grouped Likert-scale questions into *a priori*-selected topic areas to produce eight outcomes across three categories: risk (knowledge, confidence, and threat), role (efficacy and barriers), and response (ready, willing, and able). Threat and efficacy were adapted from Witte’s Extended Parallel Process Model,^16^ while other outcomes were chosen based on importance to preparedness in other occupational cohorts. Outcome variables were created by summing the responses to the respective questions within each topic area and dichotomized at the median to create a “High” and “Low” score for each outcome (details in ***Supplemental Table 2***). Some outcome topic areas combined multiple questions, while some included only one Likert-scale question, but they were treated similarly.

We performed multivariate logistic regression to evaluate associations between job role, job experience, leadership, age, gender, and contact level on each of the eight outcomes, adjusting for geographic region, race, number of dependents, living with an essential worker, and income. Associations for job role variables are presented as odds of a High score compared to a Low score within each individual job role (no reference group; e.g., odds of high knowledge within small animal veterinarians). Results for other variables are presented as comparison odds ratios; for example, the odds ratio for high scores for those with years in job greater than the median compared to less than the median. We assessed for collinearity between job and demographic variables to determine if our model variables were correlated (***Supplemental Table 3***). A final multivariate logistic regression was developed to explore the effect of risk (knowledge, confidence, and threat) and role (efficacy, barriers) as independent variables on response (ready, willing, able) outcome variables. Analysis was conducted using the R software program.^17^

## Results

### Study Population Characteristics

From 2,415 entries, a total of 1,577 (65%) participants completed all sections of the online survey and had non-missing data for key outcome variables. Statistical differences in demographics between respondents included and excluded in analysis are shown in ***Supplemental Table 4***. Participants’ job and demographic characteristics are shown in ***Table 1***. The majority (75%) of participants worked in veterinary medical clinics treating small companion animals. Within this group, most were veterinarians (38%), closely followed by veterinary technicians and assistants (32%), and other support staff (office managers, receptionists, kennel workers, etc., 5%). Eighty-three participants (5.3%) worked in other veterinary medical clinics (e.g., equine, exotic, or food animal focus), with most being veterinarians (5%). Animal rescue/control (8%), zoo/wildlife (3%), and other jobs (8%) made up the remaining participants’ job roles. The median job experience was 5 years, and 57% reported being in leadership roles. The median age was 40 years old; a detailed age distribution is included in ***Supplemental Table 1***. Most (89%) participants were female. Additional job and demographic variables are shown in ***Supplemental Table 1***.

Contact level was assessed for both co-workers and the public (clients and visitors), as shown in ***Table 2***. Participants reported more contact with co-workers than with the public. Most participants (62%) reported co-worker contact almost the entire workday, compared to public contact where the majority (43%) reported only rare daily public contact. Most participants reported interacting with 6-10 co-workers (39%) but only 1-2 clients/visitors (26%) per day. Aggregated contact quartile scores were roughly evenly distributed, ranging from 17% to 31%.

### Study Population COVID-19 Perceptions

***Table 3*** shows that most (39%) participants considered co-workers as the most likely source of SARS-CoV-2 exposure, followed by clients/visitors (32%) and the general public (25%). While participants were only able to select one response for most likely source, they were able to choose multiple answers for secondary consequences from the COVID-19 pandemic. Most participants (96%) felt there was at least one secondary consequence from the pandemic. The most frequently reported concerns were for the mental health implications for the profession (89%), concerns for the human healthcare profession (82%), and concerns for animal health and welfare (71%).

**Table 3:** Sources of and Consequences from COVID-19

|  | **N (%)** |
| --- | --- |
| **Perceived most likely source for SARS-CoV-2 exposure\*** |  |
| Co-workers | 615 (39) |
| The public as part of my job (clients / visitors) | 508 (32) |
| The general public outside of my job | 396 (25) |
| Family / friends at home | 56 (4) |
| The animals I care for | 2 (0) |
| **Perceived secondary consequences as a result of the COVID-19 pandemic\*** |  |
| Any secondary concerns | 1513 (96) |
| Mental health implications of the veterinary and animal care fields | 1399 (89) |
| Support of human healthcare professionals during and after the pandemic | 1278 (82) |
| Animal health and welfare during and after the pandemic | 1115 (71) |
| Economic resilience of the veterinary and animal care fields as a whole | 806 (51) |
| Economic resilience of your personal profession | 718 (46) |

\* Participants were able to select only one response for likely source of COVID but could choose multiple secondary consequences.

Most participants reported having no barriers to responding to work during the COVID-19 pandemic (57% for no personal barriers; 74% for no professional barriers), as shown in ***Table 4***. For those who did report barriers, the most common personal concern was for family/dependent needs (25%) and physical or mental health barriers (16% and 15%, respectively), and the most common professional concern was a lack of management support (19%).

**Table 4:** Reported Personal and Professional Barriers

|  | **N (%)** |
| --- | --- |
| **Personal Barriers** |  |
| None | 893 (57) |
| Family / dependents needs | 402 (25) |
| Health (physical) | 256 (16) |
| Health (mental) | 235 (15) |
| Transportation | 27 (2) |
| Other | 43 (3) |
| **Professional Barriers** |  |
| None | 1161 (74) |
| Lack of management support | 297 (19) |
| Lack of communication channels | 148 (9) |
| Lack of peer support | 101 (6) |
| Other | 50 (3) |

### Job and Demographics on Eight Outcomes

***Tables 5A, 5B***, and ***5C*** present the associations between job and demographic factors and the eight outcomes. There were minimal differences in the odds across job roles, as they had similar trends within outcome groups (those with higher odds were biased from a low number of participants). Participants in leadership roles had greater odds of reporting high knowledge (odds ratio (OR) 1.66 [95% confidence interval 1.37-1.94]), high confidence (OR 1.89 [1.6-2.19]), and high job efficacy (OR 2.26 [1.97-2.55]). Leadership was associated with increased odds of readiness (OR 1.33 [1.03-1.62]), willingness (OR 1.39 [1.1-1.68]), and ability (OR 1.35 [1.05-1.66]) to respond. Age also was associated with the outcomes, as participants 40 years and older had greater odds of high knowledge (OR 1.55 [1.26-1.84]), high confidence (OR 1.76 [1.46-2.06]), and high efficacy (OR 2.11 [1.81-2.41]), with lower odds of high perceived threat (OR 0.55 [0.24-0.85]) and lower odds of reported barriers (OR 0.66 [0.37-0.95]), independent of leadership role. Age correlated with response outcomes, as older participants had higher odds of readiness (OR 1.56 [1.26-1.86]), willingness (OR 1.43 [1.13-1.72]), and ability (OR 1.41 [1.1-1.72]) to respond during the COVID-19 pandemic.

**Table 5A:** Odds and Odds Ratios for Perceived Risk Outcomes

|  | **Knowledge** | **Confidence** | **Threat** |
| --- | --- | --- | --- |
|  | OR (95% Confidence Interval) | OR (95% Confidence Interval) | OR (95% Confidence Interval) |
| **Job Role - Odds** |  |  |  |
| Small Animal Medicine Veterinarian | 1.147 (0.369 - 1.925) | 0.476 (0.031 - 1.260) | 0.845 (0.035 - 1.654) |
| Small Animal Medicine Technician/Assistant | 1.078 (0.229 - 1.928) | 0.816 (0.038 - 1.671) | 0.581 (0.031 - 1.469) |
| Small Animal Medicine Support Staff | 0.697 (0.029 - 1.686) | 0.705 (0.029 - 1.700) | 0.413 (0.062 - 1.445) |
| Medicine-Other Veterinarian | 1.338 (0.443 - 2.233) | 0.707 (0.019 - 1.605) | 0.416 (0.051 - 1.338) |
| Medicine-Other Technician/Assistant | 1.072 (0.045 - 2.598) | 0.660 (0.092 - 2.242) | 0.531 (0.105 - 2.168) |
| Animal Shelter / Control | 1.156 (0.280 - 2.032) | 0.626 (0.025 - 1.505) | 0.736 (0.177 - 1.650) |
| Zoo / Wildlife | 2.454 (1.322 - 3.586) | 1.006 (0.086 - 2.097) | 0.281*(0.089 - 1.461) |
| Other Job | 1.287 (0.375 - 2.199) | 0.591 (0.032 - 1.503) | 0.392 (0.056 - 1.349) |
| **Job Experience – Odds Ratio** |  |  |  |
| Job Years greater than Median | 0.925 (0.639 - 1.211) | 0.858 (0.564 - 1.152) | 0.843 (0.541 - 1.144) |
| Job Years less than Median | *1 reference* | *1 reference* | *1 reference* |
| **Leadership – Odds Ratio** |  |  |  |
| In Leadership Role | 1.656*** (1.368 - 1.944) | 1.894*** (1.596 - 2.192) | 0.761 (0.456 - 1.066) |
| Not in Leadership Role | *1 reference* | *1 reference* | *1 reference* |
| **Age – Odds Ratio** |  |  |  |
| Age ≥ 40 years old | 1.549*** (1.258 - 1.839) | 1.760*** (1.462 - 2.058) | 0.549*** (0.244 - 0.854) |
| Age < 40 years old | *1 reference* | *1 reference* | *1 reference* |
| **Gender – Odds Ratio** |  |  |  |
| Female | 0.831 (0.329 - 1.334) | 0.714 (0.219 - 1.210) | 1.243 (0.729 - 1.757) |
| Male | *1 reference* | *1 reference* | *1 reference* |
| **Contact Level – Odds Ratio** |  |  |  |
| Co-Worker Contact Q1 | *1 reference* | *1 reference* | *1 reference* |
| Co-Worker Contact Q2 | 1.067 (0.696 - 1.437) | 0.817 (0.442 - 1.191) + | 1.878*** (1.495 - 2.262) ++ |
| Co-Worker Contact Q3 | 0.871 (0.503 - 1.239) | 0.783 (0.410 - 1.156) + | 2.187*** (1.805 - 2.569) ++ |
| Co-Worker Contact Q4 | 1.249 (0.831 - 1.666) | 0.664 (0.235 - 1.093) + | 2.664*** (2.222 - 3.105) ++ |
| Public Contact Q1 | *1 reference* | *1 reference* | *1 reference* |
| Public Contact Q2 | 1.103 (0.754 - 1.451) | 0.873 (0.519 - 1.226) + | 1.656** (1.289 - 2.023) ++ |
| Public Contact Q3 | 0.725 (0.336 - 1.115) | 0.779 (0.405 - 1.154) + | 2.100*** (1.687 - 2.512) ++ |
| Public Contact Q4 | 0.851 (0.486 - 1.216) | 0.599* (0.194 - 1.004) + | 2.123*** (1.733 - 2.513) ++ |

Controlled for job years, region, leadership, stay-at-home orders, age, gender, race, dependent number, living with an essential worker, income, and co-worker and public contact.
\* p<0.05; \*\*p<0.01; \*\*\*p<0.005
+ p-value for trend = 0.08; ++ p-value for trend < 0.005; when contact level treated as ordinal variable

**Table 5B:** Odds and Odds Ratios for Perceived Role Outcomes

|  | **Efficacy** | **Barriers** |
| --- | --- | --- |
|  | OR (95% Confidence Interval) | OR (95% Confidence Interval) |
| **Job Role - Odds** |  |  |
| Small Animal Medicine Veterinarian | 0.444 (0.037 - 1.260) | 0.592 (0.018 - 1.367) |
| Small Animal Medicine Technician/Assistant | 0.624 (0.026 - 1.512) | 0.547 (0.031 - 1.395) |
| Small Animal Medicine Support Staff | 0.639 (0.039 - 1.672) | 0.613 (0.037 - 1.598) |
| Medicine-Other Veterinarian | 0.456 (0.049 - 1.399) | 0.643 (0.024 - 1.524) |
| Medicine-Other Technician/Assistant | 0.251 (0.014 - 1.867) | 0.512 (0.012 - 2.047) |
| Animal Shelter / Control | 0.596 (0.033 - 1.523) | 0.824 (0.049 - 1.696) |
| Zoo / Wildlife | 0.637 (0.051 - 1.776) | 0.335 (0.079 - 1.460) |
| Other Job | 0.650 (0.029 - 1.598) | 0.424 (0.049 - 1.339) |
| **Job Experience – Odds Ratio** |  |  |
| Job Years greater than Median | 1.052 (0.762 - 1.342) | 0.705* (0.423 - 0.988) |
| Job Years less than Median | *1 reference* | *1 reference* |
| **Leadership – Odds Ratio** |  |  |
| In Leadership Role | 2.258*** (1.966 - 2.549) | 0.839 (0.552 - 1.126) |
| Not in Leadership Role | *1 reference* | *1 reference* |
| **Age – Odds Ratio** |  |  |
| Age ≥ 40 years old | 2.110*** (1.808 - 2.412) | 0.664** (0.374 - 0.955) |
| Age < 40 years old | *1 reference* | *1 reference* |
| **Gender – Odds Ratio** |  |  |
| Female | 0.841 (0.302 - 1.380) | 1.189 (0.690 - 1.688) |
| Male | *1 reference* | *1 reference* |
| **Contact Level – Odds Ratio** |  |  |
| Co-Worker Contact Q1 | *1 reference* | *1 reference* |
| Co-Worker Contact Q2 | 1.039 (0.654 - 1.424) | 1.015 (0.646 - 1.385) |
| Co-Worker Contact Q3 | 0.677* (0.296 - 1.057) | 1.112 (0.743 - 1.482) |
| Co-Worker Contact Q4 | 0.763 (0.332 - 1.194) | 1.094 (0.677 - 1.511) |
| Public Contact Q1 | *1 reference* | *1 reference* |
| Public Contact Q2 | 1.281 (0.921 - 1.641) | 1.179 (0.832 - 1.526) |
| Public Contact Q3 | 1.074 (0.672 - 1.475) | 0.975 (0.584 - 1.365) |
| Public Contact Q4 | 1.118 (0.743 - 1.494) | 0.927 (0.562 - 1.292) |

Controlled for job years, region, leadership, stay-at-home orders, age, gender, race, dependent number, living with an essential worker, income, and co-worker and public contact.
\* p<0.05; \*\*p<0.01; \*\*\*p<0.005, + p-value for ordinal trend <0.05, ++ p-value for trend < 0.005

**Table 5C:** Odds and Odds Ratios for Perceived Response Outcomes

|  | **Ready** | **Willing** | **Able** |
| --- | --- | --- | --- |
|  | OR (95% Confidence Interval) | OR (95% Confidence Interval) | OR (95% Confidence Interval) |
| **Job Role – Odds** |  |  |  |
| Small Animal Medicine Veterinarian | 1.071 (0.278 - 1.863) | 1.267 (0.482 - 2.052) | 1.964 (1.144 - 2.785) |
| Small Animal Medicine Technician/Assistant | 1.610 (0.740 - 2.480) | 1.765 (0.905 - 2.625) | 2.734*(1.831 - 3.636) |
| Small Animal Medicine Support Staff | 1.373 (0.355 - 2.392) | 1.835 (0.826 - 2.844) | 2.686 (1.624 - 3.748) |
| Medicine-Other Veterinarian | 1.940 (0.999 - 2.880) | 2.349 (1.419 - 3.280) | 3.920** (2.923 - 4.917) |
| Medicine-Other Technician/Assistant | 2.156 (0.525 - 3.786) | 2.553 (0.940 - 4.166) | 4.608 (2.804 - 6.412) |
| Animal Shelter / Control | 1.817 (0.898 - 2.735) | 2.006 (1.104 - 2.908) | 2.826*(1.883 - 3.769) |
| Zoo / Wildlife | 2.392 (1.195 - 3.590) | 3.174 (1.986 - 4.361) | 5.664*(4.336 - 6.993) |
| Other Job | 1.649 (0.709 - 2.590) | 1.960 (1.029 - 2.891) | 3.269*(2.278 - 4.260) |
| **Job Experience – Odds Ratio** |  |  |  |
| Job Years more than Median | 1.026 (0.733 - 1.319) | 1.069 (0.781 - 1.357) | 0.964 (0.659 - 1.268) |
| Job Years less than Median | *1 reference* | *1 reference* | *1 reference* |
| **Leadership – Odds Ratio** |  |  |  |
| In Leadership Role | 1.325 (1.031 - 1.619) | 1.387* (1.098 - 1.676) | 1.352 (1.046 - 1.658) |
| Not in Leadership Role | *1 reference* | *1 reference* | *1 reference* |
| **Age – Odds Ratio** |  |  |  |
| Age ≥40 years old | 1.563*** (1.262 - 1.865) | 1.428* (1.131 - 1.725) | 1.410* (1.097 - 1.723) |
| Age < 40 years old | *1 reference* | *1 reference* | *1 reference* |
| **Gender – Odds Ratio** |  |  |  |
| Female | 1.150 (0.634 - 1.665) | 1.042 (0.530 - 1.554) | 1.135 (0.601 - 1.670) |
| Male | *1 reference* | *1 reference* | *1 reference* |
| **Contact Level – Odds Ratio** |  |  |  |
| Co-Worker Contact Q1 | *1 reference* | *1 reference* | *1 reference* |
| Co-Worker Contact Q2 | 1.003 (0.618 - 1.388) | 0.994 (0.614 - 1.374) | 1.074 (0.673 - 1.474) |
| Co-Worker Contact Q3 | 0.702 (0.325 - 1.080) | 0.719 (0.345 - 1.093) | 0.754 (0.362 - 1.146) |
| Co-Worker Contact Q4 | 1.054 (0.620 - 1.487) | 1.069 (0.641 - 1.497) | 1.190 (0.734 - 1.645) |
| Public Contact Q1 | *1 reference* | *1 reference* | *1 reference* |
| Public Contact Q2 | 0.987 (0.626 - 1.348) | 0.951 (0.598 - 1.304) | 1.030 (0.656 - 1.403) |
| Public Contact Q3 | 1.020 (0.619 - 1.421) | 1.001 (0.606 - 1.395) | 0.956 (0.542 - 1.371) |
| Public Contact Q4 | 1.019 (0.646 - 1.392) | 1.209 (0.837 - 1.582) | 1.113 (0.722 - 1.505) |

Controlled for job years, region, leadership, stay-at-home orders, age, gender, race, dependent number, living with an essential worker, income, and co-worker and public contact.
\* p<0.05; \*\*p<0.01; \*\*\*p<0.005

The effect of contact level, with both co-workers and the public, was independently associated with the perceived threat outcome. Compared to participants in the lowest contact quartile, those with higher co-worker contact had greater odds of reporting high threat, with a dose-response increase (Q2 OR 1.88 [1.49-2.26], Q3 OR 2.18 [1.81-2.57], Q4 OR 2.66 [2.22-3.11], p-value for trend <0.005), controlling for other job and demographic variables. The same dose-response association was also observed for higher public contact with increased perceived threat (Q2 OR 1.66 [1.29-2.02], Q3 OR 2.1 [1.69-2.51], Q4 OR 2.12 [1.73-2.51], p-value for trend <0.005). Higher contact level was slightly associated with reduced odds of confidence in safety protocols, although not statistically significant (p-value for trend 0.08 for both co-workers and the public), and did not correlate with knowledge, role outcomes (job efficacy, barriers), or response outcomes (ready, willing, able).

### Perceived Risk and Role on Response Outcomes

The associations of knowledge, confidence, threat, efficacy, and barriers on the odds of high ready, willing, and able scores (response outcomes) were examined, as shown in ***Table 6***. Job efficacy had the most substantial positive correlation with response outcomes, with participants who reported high efficacy having increased odds of higher response outcomes, compared to those who reported low efficacy (Ready OR 2.82 [2.58-3.06]; Willing OR 2.34 [2.1-2.57], Able OR 2.22 [1.98-2.47]). The same association was observed in those with high confidence (Ready OR 1.94 [1.67-2.2], Willing OR 2.05 [1.79-2.31], Able OR 1.53 [1.25-1.8]). Participants who reported having more barriers had lower odds of response (Ready OR 0.28 [0.04-0.52], Willing OR 0.31 [0.08-0.54], Able OR 0.25 [0-0.49]). Perceived threat correlated with decreased willingness to respond (OR 0.72 [0.47-0.97]) but was not associated with other response outcomes. Knowledge did not significantly correlate with response outcomes.

**Table 6:** Odds Ratios for Perceived Risk and Role on Response Outcomes

|  | **Ready** | **Willing** | **Able** |
| --- | --- | --- | --- |
|  | OR (95% Confidence Interval) | OR (95% Confidence Interval) | OR (95% Confidence Interval) |
| **Knowledge** | 1.195 (0.954 - 1.436) | 1.063 (0.826 - 1.301) | 1.029 (0.783 - 1.275) |
| **Confidence** | 1.935*** (1.668 - 2.203) | 2.049*** (1.787 - 2.312) | 1.527*** (1.252 - 1.802) |
| **Threat** | 0.851 (0.597 - 1.105) | 0.721** (0.472 - 0.969) | 0.954 (0.694 - 1.214) |
| **Efficacy** | 2.822*** (2.582 - 3.061) | 2.337*** (2.101 - 2.573) | 2.222*** (1.976 - 2.468) |
| **Barriers** | 0.282*** (0.044 - 0.520) | 0.309*** (0.076 - 0.542) | 0.245*** (0.003 - 0.493) |

Low levels of Knowledge, Confidence, Perceived Threat, Efficacy, and Barriers as reference
\* p<0.05; \*\*p<0.01; \*\*\*p<0.005

## Discussion

In this study, we evaluated perceived risks and roles during the COVID-19 pandemic among VACW and found that leadership and older age had the strongest associations, independent of each other, with decreased perceived risk and increased job efficacy. We observed that increased reported contact level with others (both co-workers and the public) was associated with increased perceived risk. We further explored barriers and facilitators to VACWs’ readiness, willingness, and ability to respond during a pandemic crisis. Increased co-worker and public contact were associated with higher odds of perceived threat, yet increased contact was not correlated with response outcomes. Response outcomes were impacted by older age and leadership, as both were associated with improved readiness, willingness, and ability to respond, with lowered odds of reported barriers to response. These findings can be used to target intervention and training efforts to support this critical workforce, resulting in improved preparedness as the pandemic progresses.

Our survey data indicated that participants considered co-workers and clients/visitors their most likely source of SARS-CoV-2 exposure, even more than the general public. This finding reinforces the concept that occupational exposures are essential to consider in the context of infectious disease risk, including for COVID-19.^18^ Curiously, a low percentage of participants felt that family and friends were a significant source of SARS-CoV-2 exposure, yet data from contact tracing shows that gatherings of friends and families from separate households are a significant risk factor for COVID-19.^19^ Appropriate information on risk factors at home, and strategies to reduce transmission, based on up-to-date epidemiological data, should be included in workplace training and support plans, as this will aid in minimizing occupational exposure and spread. While it is possible that this workforce underestimates the potential risk from personal sources of exposure, it is also possible that this workforce—which includes professionals who are both well-trained in infectious disease management and are exposed to zoonotic pathogens routinely at work^3,20–22^—may use extensive measures to manage these exposures. It may also be the case that individuals feel they have more control over their exposures at home or in public (e.g., they can elect to avoid crowds), while occupational exposures may be unavoidable, or even a necessary part of their job function, and is outside of their control.

Reported contact level, both with co-workers and the public (visitors and clients), was positively associated with increased perceived threat of COVID-19, as well as a slight decrease in the confidence in safety protocols used for COVID-19. This association increased in a dose-response fashion, where every increase in contact quartile group was associated with increased perceived threat and decreased confidence. Contact level with co-workers or the public is less frequently evaluated or reported in studies on preparedness in occupational cohorts. Previous research in other occupations, such as human medicine, first responders, and public health workers, has assessed variables that are related to contact with others, such as hours worked per week,^8^ employment status (full or part time),^10,11^ shift (day/night),^10,11^ hospital size,^11^ or position/role (e.g., nurse versus physician).^10,12,13^ Nonetheless, no studies to our knowledge explicitly depict the association of self-reported contact level, either overall or divided by co-workers and the public, on response outcomes. At the same time, what is most noteworthy is that, while contact level was correlated with perceived risk, it was not correlated with response outcomes (ready, willing, and able). Participants reporting higher contact levels, both with co-workers and with the public, had higher odds of reporting increased perceived threat from COVID-19, yet they did not report that this would make them less willing, ready, or able to respond during the pandemic.

This remarkable finding demonstrates that VACW may accept even high-risk scenarios in order to deliver animal health and public health services. It is possible that this workforce is acclimated to risks due to their higher risk for exposure to zoonotic diseases.^20–22^ Nonetheless, VACWs’ high levels of response outcomes, regardless of threats, have intriguing implications for those designing public health interventions, who may otherwise overlook VACW as a valuable resource to aid in comprehensive response efforts, as our research highlights their commitment and perseverance towards public health. VACW have already been shown to enhance community pandemic response, not just through their work in infection control and expert consultation, but also through involvement in human health programs.^4,5^ Furthermore, this dedication will be particularly meaningful if the SARS-CoV-2 pathogen’s zoonotic and/or anthropozoonotic potential, i.e. its ability to transmit between humans and animals, increases as the pandemic progresses. If a new variant emerges that is more transmissible among animals or that has key animal reservoirs or vectors (conditions previously shown to be important to the epidemiology of coronaviruses^23,24^), then VACW will likely be at the frontline of disease detection and risk. This has already proven to be true in COVID-19 outbreaks among Danish mink farms related to a mink-associated variant.^4^ Establishing appropriate support and communication systems now, before more zoonotic variants arise, will have widespread benefits for public health.

Additionally, we evaluated the association of knowledge, confidence, perceived threat, efficacy, and barriers on our response outcomes (ready, willing, and able) and found that job efficacy had the strongest association with positive response. The same trend has been seen in other studies evaluating preparedness.^25^ Confidence in safety protocols was also shown to be positively correlated with increased response outcomes, but to a lesser degree than efficacy. In previous studies, confidence in safety protocols was either not evaluated or was combined with an efficacy variable.^8,25^ Future disaster response studies should consider evaluating the effect of confidence in safety protocols, independent of efficacy, which can direct training programs to improve this confidence.

Identifying that job efficacy and confidence enhanced pandemic response dispositions, we evaluated individual job and demographic factors that were associated with increased job efficacy and confidence. Being in a self-reported leadership role and being 40 years and older were associated with improved knowledge, confidence, and job efficacy, as well as reduced perceived threat. Interestingly, the years working within a job did not correlate with leadership roles and older age, and did not impact risk, role, or response outcomes. Similar trends for older age associated with improved response outcomes have been shown in other occupational groups, ^8,10–12,25^ while a minority of studies have shown the opposite effect.^13^ This finding of a positive association was contradictory to our initial assumption that older participants would report increased perceived risk, given the COVID-19 pandemic disproportionately affects older individuals for severe disease outcomes.^26^ This contradiction in perception and empirical risk is fundamental to recognize in the design of support and training systems. Intervention efforts should target all age groups, not just those at risk based on epidemiologic data. Interventions that harness the leadership’s increased job efficacy and pandemic response may hold promise to both capitalize on the strengths of leadership and address gaps among non-leaders and younger workers. One such intervention is the train-the-trainer model, which uses subject-matter experts to disseminate knowledge to instructor-trainees, who then train other groups, and is an effective and efficient way of training large groups of people in a relatively brief time period.^27–30^ Training should focus beyond increasing knowledge, which, according to our findings, did not impact response outcomes, but instead work to improve efficacy (the importance of an individual’s role in overall response efforts) and confidence in applied safety protocols.

While this study is the first to evaluate pandemic preparedness in a novel, yet critical, worker population, our study does have certain limitations. Like most volunteer questionnaire study designs, our research study is at risk of recall bias (selective memory for certain experiences/information), social desirability bias (participant responses influenced by researchers’ goals), and self-selection bias (individuals who feel strongly about a topic are more likely to participate in a study). We saw a high number of respondents who did not complete all sections of the survey. It is uncertain if this is due to the design or technical aspects of the online questionnaire or to external factors (e.g., participants were interrupted while taking the survey during working hours). However, there was no significant difference in job and demographic characteristics between those who completed the survey compared to those who did not. Another limitation is that our study population may not reflect the target veterinary and animal care workforce, limiting the external generalizability of our findings, as in the case of our low racial diversity and high percentage of female participants (veterinary medical field in US estimated at 63.9% female in 2020).^31^

Our findings suggest a need for future directions in preparedness response research within this critical worker population to address two main areas. First, there is a need to understand the relationship between changes in operational practices, at the organizational and individual levels, and changes in levels of perceived threat, efficacy, and response. Practices identified as protective can be incorporated into widespread training and support programs to improve response preparedness in this workforce for COVID-19, as well as future pandemics and other disaster situations. The second area is further exploration of the secondary consequences from the pandemic among VACW. Results from this study underscore the mental health impacts from the COVID-19 pandemic in this workforce, a concept that is mirrored in the general population.^32^ Our survey captured perceptions of mental health implications for the field as a whole; future studies should evaluate mental health effects, such as stress, anxiety, and depression, in individuals and relate that to job and demographic risk factors. Although we hypothesize that those who report higher perceived risk from COVID-19 as a result of their job will also suffer higher rates of these secondary complications, such as mental health effects, this link should be explicitly evaluated in future studies.

In conclusion, our findings highlight that veterinary and animal care workers’ perceived risk, even among high-risk groups, does not impact their response to the pandemic. This dedication to public health reinforces VACW as valuable assets in comprehensive response efforts in the community, through their role in infection control, public counselling, and supporting human health efforts. Particularly, this will have important implications if the COVID-19 develops a significant zoonotic component. To better prepare for their current and possible future roles, response to COVID-19 in VACW can be improved by targeting younger workers who are not necessarily in leadership roles, and by designing support and communication programs that improve job efficacy and confidence in safety protocols. The results of this work, and future research stemming from it, can inform interventions facilitating a more resilient workforce that is better equipped to continue responding to the COVID-19 pandemic and to future crises.

## Data Availability

Survey data is not publicly available. Please contact corresponding author for information on data sharing.

## Declarations Page

## Acknowledgements

The authors would like to thank the entire COVET Study team for their valuable contributions to this project: Kaitlin Waite, Sharmaine Miller, Cody Swilley, Meghan Brino, Joyce Kwan, Indira Chakravarti, and Justin Edwards. We would also like to thank our survey participants for their cooperation. Finally, we are grateful to Caitlin Ceryes for her expert consultation.

## Authors Contribution

Study conception and design: KRD, JA, DJB, JBN, MFD

Data collection and management: KRD, KMG, PL, MFD

Data analysis: KRD, KMG, FS, CF

Manuscript drafting: KRD

Manuscript revision: KRD, KMG, FS, CF, JRF, EAB, KM, PL, JA, DJB, JBN, MFD

## Author’s Disclosure Statement

The authors have no conflicts of interest to disclose for this manuscript.

## Funding Statement

Funding for KRD is provided by a grant from the U.S. Centers for Disease Control and Prevention, National Institute for Occupational Safety and Health to the Johns Hopkins Education and Research Center for Occupational Safety and Health [T42 OH0008428], and the AKC Canine Health Foundation Clinician-Scientist Fellowship [02525-E]. MFD was supported by the National Institutes of Health [K01OD019918]. CF was supported by the Fogarty International Center of the National Institutes of Health [D43TW009343] and the University of California Global Health Institute.

## Notes

### Competing Interest Statement

The authors have declared no competing interest.

### Author Declarations

Approval for this study was received from The Johns Hopkins Bloomberg School of Public Heath Institutional Review Board (JHSPH IRB).

## References

1. Center for Systems Science and Engineering at Johns Hopkins University. COVID-19 Dashboard. https://coronavirus.jhu.edu/map.html.

2. Nusbaum KE, Rollin BE, Wohl JS. The veterinary profession’s duty of care in response to disasters and food animal emergencies. J Am Vet Med Assoc. 2007;231(2):200–202. doi:10.2460/javma.231.2.200

3. Wenzel JGW, Nusbaum KE. Disaster medicine: Veterinary expertise in biosecurity and biological risk assessment. J Am Vet Med Assoc. 2007;230(10):1476–1480. doi:10.2460/javma.230.10.1476

4. Ferri M, Lloyd-Evans M. The contribution of veterinary public health to the management of the COVID-19 pandemic from a One Health perspective. One Heal. 2021;12:100230. doi:10.1016/j.onehlt.2021.100230

5. AVMA. Veterinarians help with COVID-19 vaccine delivery. https://www.avma.org/javma-news/2021-04-15/veterinarians-help-covid-19-vaccine-delivery. Published 2021.

6. Chiang SC, Fisher HH, Bridwell ME, Trigoso SM, Rasulnia BB, Kuwabara SA. Applying the Ready, Willing, and Able Framework to Assess Agency Public Health Emergency Preparedness: The CDC Perspective. Heal Secur. 2020;18(2):75–82. doi:10.1089/hs.2019.0090

7. Kohn S, Eaton JL, Feroz S, Bainbridge AA, Hoolachan J, Barnett DJ. Personal disaster preparedness: An integrative review of the literature. Disaster Med Public Health Prep. 2012;6(3):217–231. doi:10.1001/dmp.2012.47

8. Balicer RD, Barnett DJ, Thompson CB, et al. Characterizing hospital workers’ willingness to report to duty in an influenza pandemic through threat-and efficacy-based assessment. BMC Public Health. 2010;10. doi:10.1186/1471-2458-10-436

9. McCabe OL, Barnett DJ, Taylor HG, Links JM. Ready, willing, and able: A framework for improving the public health emergency preparedness system. Disaster Med Public Health Prep. 2010;4(2):161–168. doi:10.1001/dmp-v4n2-hcn10003

10. Qureshi K, Gershon RRM, Sherman MF, et al. Health care workers’ ability and willingness to report to duty during catastrophic disasters. J Urban Heal. 2005;82(3):378–388. doi:10.1093/jurban/jti086

11. Charney RL, Rebmann T, Flood RG. Hospital Employee Willingness to Work during Earthquakes Versus Pandemics. J Emerg Med. 2015;49(5):665–674. doi:10.1016/j.jemermed.2015.07.030

12. Burke R V., Goodhue CJ, Chokshi NK, Upperman JS. Factors associated with willingness to respond to a disaster: A study of healthcare workers in a tertiary setting. Prehosp Disaster Med. 2011;26(4):244–250. doi:10.1017/S1049023X11006492

13. Ogedegbe C, Nyirenda T, DelMoro G, Yamin E, Feldman J. Health care workers and disaster preparedness: Barriers to and facilitators of willingness to respond. Int J Emerg Med. 2012;5(1):1–9. doi:10.1186/1865-1380-5-29

14. Turner JA, Rebmann T, Loux TM, Charney RL. Willingness to Respond to Radiological Disasters among First Responders in St. Louis, Missouri. Heal Secur. 2020;18(4):318–328. doi:10.1089/hs.2019.0160

15. Harris PA, Taylor R, Minor BL, et al. The REDCap consortium: Building an international community of software platform partners. J Biomed Inform. 2019;95(May):103208. doi:10.1016/j.jbi.2019.103208

16. Health Communication Capacity Collaborative. The Extended Parallel Processing Model.; 2011.

17. R Development Core Team. R: a language and environment for statistical computing. Vienna R Found Stat Comput. 2010.

18. Baker MG, Peckham TK, Seixas NS. Estimating the burden of United States workers exposed to infection or disease: A key factor in containing risk of COVID-19 infection. PLoS One. 2020;15(4):4–11. doi:10.1371/journal.pone.0232452

19. Centers for Disease Control and Prevention. Considerations for Events and Gatherings. https://www.cdc.gov/coronavirus/2019-ncov/community/large-events/considerations-for-events-gatherings.html. Published 2021.

20. Robin C, Bettridge J, McMaster F. Zoonotic disease risk perceptions in the British veterinary profession. Prev Vet Med. 2017;136:39–48. doi:10.1016/j.prevetmed.2016.11.015

21. Dowd K, Taylor M, Toribio Jalml, Hooker C, Dhand NK. Zoonotic disease risk perceptions and infection control practices of Australian veterinarians: Call for change in work culture. Prev Vet Med. 2013;111(1-2):17–24. doi:10.1016/j.prevetmed.2013.04.002

22. Williams CJ, Scheftel JM, Elchos BL, Hopkins SG, Levine JF. Compendium of Veterinary Standard Precautions for Zoonotic Disease Prevention in Veterinary Personnel: National Association of State Public Health Veterinarians: Veterinary Infection Control Committee 2015. JAVMA. 2015;247(11):1252–1277.

23. Tiwari R, Dhama K, Sharun K, et al. COVID-19: animals, veterinary and zoonotic links. Vet Q. 2020;40(1):169–182. doi:10.1080/01652176.2020.1766725

24. Davis MF, Innes GK. The Cat’s in the Bag: Despite limited cat-to-cat SARS-CoV-2 transmission, One Health surveillance efforts are needed. J Infect Dis. 2021;223 April(8):1309–1312. doi:10.1093/infdis/jiab106

25. Barnett DJ, Balicer RD, Thompson CB, et al. Assessment of local public health workers’ willingness to respond to pandemic influenza through application of the extended parallel process model. PLoS One. 2009;4(7). doi:10.1371/journal.pone.0006365

26. Centers for Disease Control and Prevention. COVID-19 and Older Adults. 2021. https://www.cdc.gov/coronavirus/2019-ncov/need-extra-precautions/older-adults.html.

27. World Health Organization. WHO Train the Trainer Workshop: Developing National Deployment and Vaccination Plans (NDVP) for Pandemic Influenza Vaccines, Geneva, Switzerland, 10–12 September 2019.; 2020.

28. Cross W, Cerulli C, Richards H, He H, Herrmann J. Predicting Dissemination of a Disaster Mental Health “Train-the-trainer” Program. Disaster Med Public Health Prep. 2010;4(4):339. doi:10.1001/dmp.2010.6.Predicting

29. Lamb MJ, La Delfa A, Sawhney M, et al. Implementation and Evaluation of an IPAC SWAT Team Mobilized to Long-Term Care and Retirement Homes During the COVID-19 Pandemic: A Pragmatic Health System Innovation. J Am Med Dir Assoc. 2021;22(2):253–255.e1. doi:10.1016/j.jamda.2020.11.033

30. Jacobs-Wingo JL, Schlegelmilch J, Berliner M, Airall-Simon G, Lang W. Emergency preparedness training for hospital nursing staff, New York City, 2012--2016. J Nurs Scholarsh. 2019;51(1):81–87.

31. AVMA. U.S. veterinarians 2020. https://www.avma.org/resources-tools/reports-statistics/market-research-statistics-us-veterinarians. Published 2021.

32. Pfefferbaum B, North CS. Mental Health and the Covid-19 Pandemic. N Engl J Med. 2020;383(6):508–510. doi:10.1056/nejmp2013466

