## Supplementary Tables for "Assessing COVID-19 Pandemic Risk Perception and Response Preparedness in Veterinary and Animal Care Workers"

Dalton et. al., 2021

#### Supplemental Table 1 - Extended Job and Demographic Characteristics

| Job & Demographic Variables N(%) | data (N = 1,577) |
|---|---|
| **US Region** | |
| Northeast/Mid-Atlantic | 515 (33) |
| Southeast | 243 (16) |
| North Midwest | 281 (18) |
| South Midwest | 133 (9) |
| Mountain | 159 (10) |
| Pacific | 186 (12) |
| Outside Continental US | 34 (2) |
| **Area in Stay-at-Home Order** | |
| Yes | 949 (60) |
| **Age** | |
| 18-29 | 274 (17) |
| 30-39 | 541 (34) |
| 40-49 | 412 (26) |
| 50-59 | 224 (14) |
| 60-69 | 103 (7) |
| 69+ | 16 (1) |
| Prefer Not to Say | 4 (0) |
| **Race/Ethnicity** | |
| White | 1,374 (87) |
| Hispanic / Latino | 66 (4) |
| Black / African American | 15 (1) |
| Asian | 34 (2) |
| Other | 82 (5) |
| **Marital Status** | |
| Married / Domestic Partnership | 958 (61) |
| Single / Divorced / Widowed | 581 (37) |
| Other / Prefer Not to Say | 29 (2) |
| **Household Number** | |
| Min | 0 |
| Mean +/- SD | 2.62 ± 1.32 |
| Median (IQR) | 2.00 (2.00, 3.00) |
| Max | 20 |
| **Dependent Number** | |
| Min | 0 |
| Mean +/- SD | 0.68 ± 1.01 |
| Median (IQR) | 0.00 (0.00, 1.00) |
| Max | 6 |
| **Living with Essential Worker** | |
| Yes | 695 (44) |
| **Annual Household Income** | |
| Less than $50,000 | 259 (17) |

| Job & Demographic Variables N(%) | data (N = 1,577) |
|---|---|
| $50,001 – $100,000 | 427 (27) |
| $100,001 - $200,000 | 561 (36) |
| Greater than $200,000 | 187 (12) |
| Prefer Not to Say | 124 (8) |

### Supplemental Table 2 - Distribution of Likert Questions and Outcome Aggregated Scores

#### Aggregated Knowledge Score

How knowledgeable are you regarding the ..... (1 = least knowledgeable, 5 = most knowledgeable)

| Label | Stats / Values | Freqs (% of Valid) | Graph | Valid |
|---|---|---|---|---|
| Public health impacts of the pandemic in general? | Mean (sd) : 3.7 (0.7)<br>min < med < max:<br>1 < 4 < 5<br>IQR (CV) : 1 (0.2) | 1 : 11 ( 0.7% )<br>2 : 67 ( 4.3% )<br>3 : 498 ( 31.6% )<br>4 : 844 ( 53.6% )<br>5 : 156 ( 9.9% ) | | 1576<br>(99.9%) |
| Your personal risk during the pandemic? | Mean (sd) : 4 (0.7)<br>min < med < max:<br>1 < 4 < 5<br>IQR (CV) : 0 (0.2) | 1 : 8 ( 0.5% )<br>2 : 33 ( 2.1% )<br>3 : 253 ( 16.1% )<br>4 : 964 ( 61.2% )<br>5 : 316 ( 20.1% ) | | 1574<br>(99.8%) |
| Ability to answer pandemic-related questions from clients or visitors? | Mean (sd) : 3.6 (0.8)<br>min < med < max:<br>1 < 4 < 5<br>IQR (CV) : 1 (0.2) | 1 : 14 ( 0.9% )<br>2 : 136 ( 8.6% )<br>3 : 449 ( 28.5% )<br>4 : 764 ( 48.6% )<br>5 : 210 ( 13.4% ) | | 1573<br>(99.7%) |
| Safety practices at work that are specific to COVID-19? | Mean (sd) : 4.1 (0.8)<br>min < med < max:<br>1 < 4 < 5<br>IQR (CV) : 1 (0.2) | 1 : 6 ( 0.4% )<br>2 : 56 ( 3.6% )<br>3 : 202 ( 12.8% )<br>4 : 870 ( 55.3% )<br>5 : 439 ( 27.9% ) | | 1573<br>(99.7%) |

#### Aggregated Confidence Score

How confident are you regarding the ..... (1 = least confident, 5 = most confident)

| Label | Stats / Values | Freqs (% of Valid) | Graph | Valid |
|---|---|---|---|---|
| Your safety at work in general? | Mean (sd) : 3.8 (0.9)<br>min < med < max:<br>1 < 4 < 5<br>IQR (CV) : 1 (0.2) | 1 : 22 ( 1.4% )<br>2 : 104 ( 6.6% )<br>3 : 399 ( 25.3% )<br>4 : 708 ( 45.0% )<br>5 : 341 ( 21.7% ) | | 1574<br>(99.8%) |
| Your safety at work that is specific to COVID-19? | Mean (sd) : 3.2 (1.2)<br>min < med < max:<br>1 < 3 < 5<br>IQR (CV) : 2 (0.4) | 1 : 132 ( 8.4% )<br>2 : 303 ( 19.3% )<br>3 : 471 ( 30.0% )<br>4 : 451 ( 28.7% )<br>5 : 215 ( 13.7% ) | | 1572<br>(99.7%) |

#### Aggregated Threat Score

What is your likelihood of ..... (1 = least likely, 5 = most likely)

| Label | Stats / Values | Freqs (% of Valid) | Graph | Valid |
|---|---|---|---|---|
| Being exposed to SARS-CoV-2 as part of your job function? | Mean (sd) : 3.1 (1.2)<br>min < med < max:<br>1 < 3 < 5<br>IQR (CV) : 2 (0.4) | 1 : 132 ( 8.4% )<br>2 : 354 ( 22.5% )<br>3 : 522 ( 33.1% )<br>4 : 342 ( 21.7% )<br>5 : 226 ( 14.3% ) | | 1576<br>(99.9%) |

| Label | Stats / Values | Freqs (% of Valid) | Graph | Valid |
|---|---|---|---|---|
| You becoming sick with COVID-19 disease as a result of your job? | Mean (sd) : 2.8 (1.1)<br>min < med < max:<br>1 < 3 < 5<br>IQR (CV) : 2 (0.4) | 1 : 197 ( 12.5% )<br>2 : 430 ( 27.3% )<br>3 : 528 ( 33.5% )<br>4 : 282 ( 17.9% )<br>5 : 139 ( 8.8% ) | | 1576<br>(99.9%) |
| Your co-workers becoming sick with COVID-19 as a result of their job? | Mean (sd) : 3 (1.2)<br>min < med < max:<br>1 < 3 < 5<br>IQR (CV) : 2 (0.4) | 1 : 158 ( 10.1% )<br>2 : 381 ( 24.3% )<br>3 : 505 ( 32.1% )<br>4 : 331 ( 21.1% )<br>5 : 196 ( 12.5% ) | | 1571<br>(99.6%) |
| Becoming seriously ill with COVID-19 disease as a result of your job? | Mean (sd) : 3.3 (1.2)<br>min < med < max:<br>1 < 3 < 5<br>IQR (CV) : 2 (0.4) | 1 : 141 ( 9.0% )<br>2 : 315 ( 20.1% )<br>3 : 414 ( 26.4% )<br>4 : 400 ( 25.5% )<br>5 : 296 ( 18.9% ) | | 1566<br>(99.3%) |
| You spreading SARS-CoV-2 to others around you? | Mean (sd) : 2.6 (1.2)<br>min < med < max:<br>1 < 3 < 5<br>IQR (CV) : 1 (0.4) | 1 : 282 ( 18.0% )<br>2 : 476 ( 30.4% )<br>3 : 447 ( 28.5% )<br>4 : 253 ( 16.1% )<br>5 : 110 ( 7.0% ) | | 1568<br>(99.4%) |

Do you agree with the following statements .... (1 = strongly disagree, 5 = strongly agree)

| Label | Stats / Values | Freqs (% of Valid) | Graph | Valid |
|---|---|---|---|---|
| The COVID-19 pandemic has severe public health consequences to those around me. | Mean (sd) : 4.1 (1)<br>min < med < max:<br>1 < 4 < 5<br>IQR (CV) : 1 (0.3) | 1 : 39 ( 2.5% )<br>2 : 89 ( 5.6% )<br>3 : 259 ( 16.4% )<br>4 : 436 ( 27.6% )<br>5 : 754 ( 47.8% ) | | 1577<br>(100.0%) |
| The COVID-19 pandemic is likely to negatively influence my job role. | Mean (sd) : 3.1 (1.3)<br>min < med < max:<br>1 < 3 < 5<br>IQR (CV) : 2 (0.4) | 1 : 242 ( 15.3% )<br>2 : 326 ( 20.7% )<br>3 : 374 ( 23.7% )<br>4 : 373 ( 23.7% )<br>5 : 262 ( 16.6% ) | | 1577<br>(100.0%) |
| I am at risk of COVID-19 disease because of my job role | Mean (sd) : 3.6 (1.2)<br>min < med < max:<br>1 < 4 < 5<br>IQR (CV) : 1 (0.3) | 1 : 109 ( 6.9% )<br>2 : 203 ( 12.9% )<br>3 : 281 ( 17.8% )<br>4 : 590 ( 37.4% )<br>5 : 394 ( 25.0% ) | | 1577<br>(100.0%) |
| Others around me are at risk of COVID-19 disease because of my job role. | Mean (sd) : 3.4 (1.3)<br>min < med < max:<br>1 < 4 < 5<br>IQR (CV) : 2 (0.4) | 1 : 169 ( 10.7% )<br>2 : 233 ( 14.8% )<br>3 : 308 ( 19.5% )<br>4 : 523 ( 33.2% )<br>5 : 344 ( 21.8% ) | | 1577<br>(100.0%) |

#### Aggregated Efficacy Score

What is the importance of ..... (1 = least important, 5 = most important)

| Label | Stats / Values | Freqs (% of Valid) | Graph | Valid |
|---|---|---|---|---|
| Your job roles and responsibilities during a pandemic or other emergent situation? | Mean (sd) : 4.1 (0.9)<br>min < med < max:<br>1 < 4 < 5<br>IQR (CV) : 1 (0.2) | 1 : 18 ( 1.1% )<br>2 : 75 ( 4.8% )<br>3 : 266 ( 17.0% )<br>4 : 562 ( 35.8% )<br>5 : 647 ( 41.3% ) | | 1568<br>(99.4%) |
| Your individual role to the overall function of your workplace during a pandemic or other emergent situation? | Mean (sd) : 4.3 (0.9)<br>min < med < max:<br>1 < 5 < 5<br>IQR (CV) : 1 (0.2) | 1 : 22 ( 1.4% )<br>2 : 44 ( 2.8% )<br>3 : 211 ( 13.4% )<br>4 : 505 ( 32.2% )<br>5 : 788 ( 50.2% ) | | 1570<br>(99.6%) |

Do you agree with the following statements .... (1 = strongly disagree, 5 = strongly agree)

| Label | Stats / Values | Freqs (% of Valid) | Graph | Valid |
|---|---|---|---|---|
| I am able to perform my duties successfully during the COVID-19 pandemic. | Mean (sd) : 4.2 (0.9)<br>min < med < max:<br>1 < 4 < 5<br>IQR (CV) : 1 (0.2) | 1 : 26 ( 1.6% )<br>2 : 85 ( 5.4% )<br>3 : 143 ( 9.1% )<br>4 : 658 ( 41.7% )<br>5 : 665 ( 42.2% ) | | 1577<br>(100.0%) |
| My co-workers are able to perform their duties successfully during the COVID-19 pandemic. | Mean (sd) : 4.1 (0.9)<br>min < med < max:<br>1 < 4 < 5<br>IQR (CV) : 1 (0.2) | 1 : 21 ( 1.3% )<br>2 : 115 ( 7.3% )<br>3 : 179 ( 11.4% )<br>4 : 692 ( 43.9% )<br>5 : 570 ( 36.1% ) | | 1577<br>(100.0%) |
| If I perform my duties successfully, it will make a big difference in the success of the response to the COVID-19 pandemic. | Mean (sd) : 3.2 (1.2)<br>min < med < max:<br>1 < 3 < 5<br>IQR (CV) : 2 (0.4) | 1 : 169 ( 10.7% )<br>2 : 248 ( 15.7% )<br>3 : 496 ( 31.5% )<br>4 : 372 ( 23.6% )<br>5 : 292 ( 18.5% ) | | 1577<br>(100.0%) |

#### Aggregated Barriers Score

What is your likelihood of ..... (1 = least likely, 5 = most likely)

| Label | Stats / Values | Freqs (% of Valid) | Graph | Valid |
|---|---|---|---|---|
| Have personal barriers that prevent you from reporting to work on-site? | Mean (sd) : 1.8 (1.1)<br>min < med < max:<br>1 < 1 < 5<br>IQR (CV) : 1 (0.6) | 1 : 896 ( 56.9% )<br>2 : 364 ( 23.1% )<br>3 : 176 ( 11.2% )<br>4 : 76 ( 4.8% )<br>5 : 62 ( 3.9% ) | | 1574<br>(99.8%) |
| Have professional barriers that prevent you from reporting to work on-site? | Mean (sd) : 1.5 (0.9)<br>min < med < max:<br>1 < 1 < 5<br>IQR (CV) : 1 (0.6) | 1 : 1107 ( 70.6% )<br>2 : 286 ( 18.2% )<br>3 : 102 ( 6.5% )<br>4 : 39 ( 2.5% )<br>5 : 34 ( 2.2% ) | | 1568<br>(99.4%) |

#### Aggregated Response Outcome Score

Each question aggregated separately to create the three response outcomes (ready, willing, able)

Do you agree with the following statements .... (1 = strongly disagree, 5 = strongly agree)

| Label | Stats / Values | Freqs (% of Valid) | Graph | Valid |
|---|---|---|---|---|
| I am prepared and ready to respond to work during the COVID-19 pandemic. | Mean (sd) : 4.5 (0.8)<br>min < med < max:<br>1 < 5 < 5<br>IQR (CV) : 1 (0.2) | 1 : 20 ( 1.3% )<br>2 : 41 ( 2.6% )<br>3 : 117 ( 7.5% )<br>4 : 383 ( 24.6% )<br>5 : 996 ( 64.0% ) | | 1557<br>(98.7%) |
| I am willing to respond to work during the COVID-19 pandemic. | Mean (sd) : 4.5 (0.8)<br>min < med < max:<br>1 < 5 < 5<br>IQR (CV) : 1 (0.2) | 1 : 15 ( 1.0% )<br>2 : 35 ( 2.2% )<br>3 : 114 ( 7.3% )<br>4 : 412 ( 26.3% )<br>5 : 992 ( 63.3% ) | | 1568<br>(99.4%) |
| I feel I am able to respond to work during the COVID-19 pandemic. | Mean (sd) : 4.6 (0.7)<br>min < med < max:<br>1 < 5 < 5<br>IQR (CV) : 1 (0.2) | 1 : 11 ( 0.7% )<br>2 : 20 ( 1.3% )<br>3 : 74 ( 4.7% )<br>4 : 372 ( 23.7% )<br>5 : 1090 ( 69.6% ) | | 1567<br>(99.4%) |

#### Distribution of Aggregated Outcome Scores

| Label | Stats / Values | Freqs (% of Valid) | Graph | Valid |
|---|---|---|---|---|

| Label | Stats / Values | Freqs (% of Valid) | Graph | Valid |
|---|---|---|---|---|
| Aggregated Knowledge Score | 1. Low<br>2. High | 737 (47.1%)<br>828 (52.9%) | | 1565<br>(99.2%) |
| Aggregated Confidence Score | 1. Low<br>2. High | 906 (57.6%)<br>666 (42.4%) | | 1572<br>(99.7%) |
| Aggregated Threat Score | 1. Low<br>2. High | 721 (46.5%)<br>829 (53.5%) | | 1550<br>(98.3%) |
| Aggregated Efficacy Score | 1. Low<br>2. High | 656 (42.0%)<br>906 (58.0%) | | 1562<br>(99.0%) |
| Aggregated Barriers Score | 1. Low<br>2. High | 842 (53.8%)<br>724 (46.2%) | | 1566<br>(99.3%) |
| Aggregated Readiness Score | 1. Low<br>2. High | 561 (36.0%)<br>996 (64.0%) | | 1557<br>(98.7%) |
| Aggregated Willingness Score | 1. Low<br>2. High | 576 (36.7%)<br>992 (63.3%) | | 1568<br>(99.4%) |
| Aggregated Ability Score | 1. Low<br>2. High | 477 (30.4%)<br>1090 (69.6%) | | 1567<br>(99.4%) |

### Supplemental Table 3 - Correlation Matrixes to test for Collinearity

job.n = job role, job_time_binary.n = time in job, job_region = location, job_home = stay-at-home orders, dem_age = age, dem_gender = gender, dem_race = race/ethnicity, dem_marital = martial status, dem_housenum = household number, dem_dependnum = number of household dependents, dem_essenital = living with an essential worker, dem_salary1 = annual salary

**Correlation Matrix**

| | job.n | job_time_binary.n | job_region | job_leader | job_home | dem_age | dem_gender | dem_race | dem_marital | dem_housenum | dem_dependnum | dem_essential1 | den |
|---|---|---|---|---|---|---|---|---|---|---|---|---|---|
| job.n | 1 | 0.042 | 0.072 | -0.021 | -0.051 | 0.037 | 0.023 | -0.048 | 0.025 | 0.060 | 0.100 | -0.031 | |
| job_time_binary.n | 0.042 | 1 | 0.003 | 0.282 | -0.012 | 0.424 | -0.131 | 0.031 | 0.139 | 0.014 | 0.093 | -0.043 | |
| job_region | 0.072 | 0.003 | 1 | 0.031 | 0.095 | 0.035 | -0.009 | -0.014 | 0.020 | 0.023 | 0.031 | -0.004 | |
| job_leader | -0.021 | 0.282 | 0.031 | 1 | -0.043 | 0.270 | -0.095 | 0.030 | 0.094 | 0.086 | 0.139 | 0.049 | |
| job_home | -0.051 | -0.012 | 0.095 | -0.043 | 1 | 0.003 | 0.078 | 0.034 | -0.010 | -0.059 | -0.091 | 0.001 | |
| dem_age | 0.037 | 0.424 | 0.035 | 0.270 | 0.003 | 1 | -0.228 | 0.037 | 0.287 | 0.005 | 0.096 | -0.112 | |
| dem_gender | 0.023 | -0.131 | -0.009 | -0.095 | 0.078 | -0.228 | 1 | 0.090 | -0.032 | 0.001 | -0.032 | 0.004 | - |
| dem_race | -0.048 | 0.031 | -0.014 | 0.030 | 0.034 | 0.037 | 0.090 | 1 | 0.073 | -0.023 | -0.023 | -0.064 | |
| dem_marital | 0.025 | 0.139 | 0.020 | 0.094 | -0.010 | 0.287 | -0.032 | 0.073 | 1 | 0.239 | 0.252 | 0.104 | |
| dem_housenum | 0.060 | 0.014 | 0.023 | 0.086 | -0.059 | 0.005 | 0.001 | -0.023 | 0.239 | 1 | 0.781 | 0.289 | |
| dem_dependnum | 0.100 | 0.093 | 0.031 | 0.139 | -0.091 | 0.096 | -0.032 | -0.023 | 0.252 | 0.781 | 1 | 0.132 | |
| dem_essential1 | -0.031 | -0.043 | -0.004 | 0.049 | 0.001 | -0.112 | 0.004 | -0.064 | 0.104 | 0.289 | 0.132 | 1 | |
| dem_salary1 | 0.197 | 0.140 | 0.043 | 0.165 | 0.015 | 0.315 | -0.126 | 0.016 | 0.174 | 0.241 | 0.188 | 0.100 | |

No signficance based on Spearman correlation statistic

### Supplemental Table 4 - Chi-Squared Comparison in Key Variables between Complete and Non-Complete Survey Respondents

Differences in job and demographic factors between respondents who completed all sections of the survey and those who did not

| Variable | Chi-Squared p-value |
|---|---|
| Job Role | 0.002 |
| Time in Job | 0.4298 |
| Leadership Role | 0.1589 |
| Age | 0.001 |
| Gender | 0.2004 |
| US Region | 0.5612 |
| Area in Stay-at-Home Order | 0.2529 |
| Race | 0.9595 |
| Marital Status | 0.5752 |
| Dependent Number | 0.2469 |
| Living with an Essential Worker | 0.933 |
| Annual Household Income | 0.4593 |

.
